# Short report: Targeted analysis of whole exome sequencing data in Indian cryptogenic stroke patients

**DOI:** 10.1101/2025.06.03.25328938

**Authors:** Priya Dev, Jenefer M. Blackwell, Rajiv Kumar, Vijay Nath Mishra, Abhishek Pathak

## Abstract

Cryptogenic stroke (CS) is an ischemic stroke of unknown cause with increasing incidence in India. Common and rare genetic variants have been associated with the risk of stroke. We carried out targeted analysis of whole exome sequencing on a small cohort of 16 CS patients to determine whether rare coding variants in genes previously associated with stroke could play a role in India. Variants were filtered for coverage (≥15x) and minor allele frequency (≤0.01). Potential pathogenic variants were classified according to ACMG guidelines, and targeted analysis performed by filtering for these variants in a panel of 220 stroke-related genes. Enrichr and STRING were used to look for enrichment of gene sets and pathways in putative deleterious (PHRED-scaled CADD scores ≥15) variants identified in CS cases. One pathogenic and 26 variants of uncertain significance were identified in 19 genes in CS patients. Enrichr showed significant enrichment for gene-sets in phenotypes (e.g. hemorrhage; abnormal blood coagulation) and pathways (e.g. common pathway of fibrin clot formation; response to elevated platelet cytosolic Ca^2+^) that could contribute to CS, indicating specific genes (e.g. *IGA2B, F13A1, F5, ATP7A, JAG1, PKD1, PMM2, GLA*) to be targeted for follow-up.

## Introduction

Stroke is the second largest cause of death and the third largest cause of years of life lost worldwide. Ischemic stroke, an overt symptomatic expression of brain infarction, accounts for ∼80% of all strokes, with most cases caused by a combination of environmental and genetic factors. Cryptogenic (unexplained) stroke (CS) accounts for ∼30%–40% of ischemic stroke patients, and is increasing in the Indian population [1]. A better definition and identification of associated risk factors are required to manage CS.

Heritability for ischemic stroke is substantial (37.9%) and varies with subtype (40.3% for large-vessel disease; 32.6% for cardioembolic; and 16.1% for small vessel disease) [2]. Genome-wide studies have identified common variants in a large number of genes associated with stroke, including in India [3]. Rare genetic variants in monogenic disorders can also lead to ischemic stroke (OMIM#601367). Evidence is accumulating for the contribution of rare functional coding variants to genetic risk in complex diseases [4], while next-generation sequencing has made finding these more cost-effective. Here we employ targeted analysis of whole exome sequencing (WES) data in Indian CS patients to determine whether rare putative deleterious variants in previously identified stroke genes could play a role in disease risk.

## Materials and Methods

All procedures in this study were conducted according to the principles of the Declaration of Helsinki. This study was approved by Institutional Ethics Committee, IMS, BHU, Varanasi, India with reference number: Dean/2018/EC/288. Patients were enrolled between 26 August 2020 and 30 March 2022. Written consent for participation was obtained from all the patients/persons responsible. All patients whose DNA was collected for this research consented to storage of the sample and future use of de-identified genetic and clinical data. All participants agreed to publication of de-identified genetic and clinical data.

### Study subjects

The study was conducted at the Department of Neurology, Institute of Medical Sciences, Sir Sunderlal Hospital, Banaras Hindu University, Varanasi, India. We enrolled 16 consecutive CS patients (age range 47-84 years; 10 males, 6 females) with ischemic strokes of undetermined etiology. CS was defined as an ischemic stroke not attributed to a definite source of large-vessel atherosclerosis, cardioembolism, or small vessel disease, according to the Trial of ORG 10172 in Acute Stroke Treatment (TOAST) classification [5]. Patients presenting within 48h of onset of clinical symptoms and with a modified Rankin Scale (mRS) score ≥1 at admission were included. Complete evaluation, including clinical assessment and neurological examination, was carried out in line with the National Institutes of Health Stroke Scale within 60 min of the patient’s arrival. The diagnostic assessment included non-contrast computed tomography or brain magnetic resonance imaging, hemogram, biochemical tests, electrocardiogram, transoesophageal echocardiography, vascular imaging (of intra- and extra-cranial vessels), assessment of prothrombotic state, and 24h Holter monitoring for atrial fibrillation. Patients with conditions requiring intensive care unit management, pregnancy, recurrent ischemic stroke, subarachnoid hemorrhage, traumatic brain injury, vascular aneurysm, arterial malformation, infective endocarditis, central nervous system infections or chronic liver or kidney diseases were excluded.

### Library preparation and exome sequencing

DNA was extracted from 2 ml of peripheral blood using Qiagen DNA mini kits, with quantity/quality of genomic DNA (gDNA) measured by NanoDrop-2000 Spectrophotometer. DNA samples were sent to Dr Lal PathLabs Ltd (New Delhi, India) where library construction, sequencing and data analyses were undertaken. Libraries were constructed from 100ng gDNA using the Ion Ampliseq Exome RDY Panel kit (Thermo Fisher Scientific) and quantified with Qubit™ dsDNA HS (High Sensitivity) Assay Kit on Qubit 3.0 Fluorometer. Templates were generated using 25pm of each library (Ion Chef Instrument; Thermo Fisher Scientific), followed by enrichment of templated ion sphere particles. WES was performed using Hi-Q chemistry on the Ion Proton system (Thermo Fisher Scientific).

### Data processing and variant analysis

Sequences were aligned against the reference genome (GRCh37/hg19) using Torrent Suite v.5.12.0 and Variant Caller v.5.2.1 software, including coverage analysis and variant caller plugins (Thermo Fisher Scientific). Variant discovery, genotype calling of multi-allelic substitutions and indels were performed using the Torrent Variant Caller version 4.6.0.7. The Torrent Coverage Analysis provided statistics and graphs describing the level of sequence coverage produced for targeted genomic regions (version 4.6.0.3). The Annotate variants 5.0 of Ion Reporter (Thermo Fisher) annotated the variants.

### Variant prioritization and bioinformatics analysis

An Integrative Genome Viewer (https://www.broadinstitute.org/igv/) was used to visualise sequencing data. Variant frequencies were obtained from public domain databases including the 1000 Genomes Project (https://www.internationalgenome.org/) and the Genome Aggregation Database (gnomAD; https://gnomad.broadinstitute.org/). Variants detected in the exome sequencing were filtered for coverage (≥15x) and minor allele frequency (≤0.01) in the public domain databases. All were compared with mutation databases including the Human Gene Mutation Database; https://www.hgmd.cf.ac.uk/ac/index.php/) and Uniprot (https://www.uniprot.org/). Intronic, up/downstream, and synonymous variants were removed. Potential pathogenicity of the detected variants was evaluated using Mutation Taster (https://www.mutationtaster.org/), Sorting Intolerant From Tolerant (SIFT; https://sift.bii.a-star.edu.sg/), Protein Variant Effect Analyzer (https://provean.jcvi.org/index.php), Functional Analysis Through Hidden Markov Model (https://fathmm.biocompute.org.uk/) and Deleterious Annotation of Genetic Variation using Neural Networks (Index of /public_data/DANN). Variants were classified as pathogenic, likely pathogenic, variant of uncertain signficiance (VUS), likely benign, and benign according to American College of Medical Genetics and Genomics (ACMG) 2015 guidelines.

### Targeted exome analysis

For targeted analysis variants classified as pathogenic, likely pathogenic, or VUS were filtered against an in-house gene panel (Dr Lal Pathlabs Ltd) of 220 genes (S1 Table) previously identified as risk factors for stroke and related clinical phenotypes. PHRED-scaled Combined Annotation Dependent Depletion (CADD) scores[6] were determined for all variants in these genes. Although there is no hard cut-off to determine potential pathogenicity in VUS, a cut-off of 15 for scaled CADD scores has been suggested [6]. All variants were examined in ClinVar (https://www.ncbi.nlm.nih.gov/clinvar/), the public archive of interpretations of clinically relevant variants. Genes with variants in CS patients were analysed using the knowledge base of known and predicted protein-protein interactions STRING database (https://string-db.org/). Gene-sets with variants of potential pathogenicity were identified using the comprehensive gene set enrichment tool EnrichR (https://maayanlab.cloud/Enrichr/).

## Results

### The study cohort

S2 Table shows clinical data and family history of stroke-related diseases for the 16 CS patients (designated P1 to P16). All except P1 and P16 presented with hypertension; 4 (P3, P11, P12, P13) had diabetes; all had Fazekas scores of 2 or 3 for white matter hyperintensities. All except P10 had a family history of stroke; all except P1, P9 and P10 reported family history of hypertension. Five patients (P2, P3, P4, P7, P11) reported a history of diabetes in a first degree relative; 2 CS patients (P12, P13) had first degree relatives who had suffered heart attacks with myocardial infarctions.

### Genetic variants identified

In all, 26 VUS in 19 genes and one asymptomatic heterozygous pathogenic carrier at autosomal recessive *PMM2* were identified (Table 1). P2, P5 and P16 had no putative deleterious variants in the targeted panel. Of the 26 VUS, 23 were heterozygous at autosomal genes, one homozygous autosomal variant at *ITGA2B*, and two hemizygous X-linked VUS at *ATP7A* and at *GLA*. Although pathogenic variants at these three genes cause serious monogenic disorders, these CS patients did not present with clinical signs relevant to them. All VUS in CS patients had PHRED-scaled CADD scores ≥15 or were disruptive in-frame deletions and hence have the potential to be deleterious. The 19 genes with putative deleterious variants were taken forward in downstream analyses.

**Table 1.**
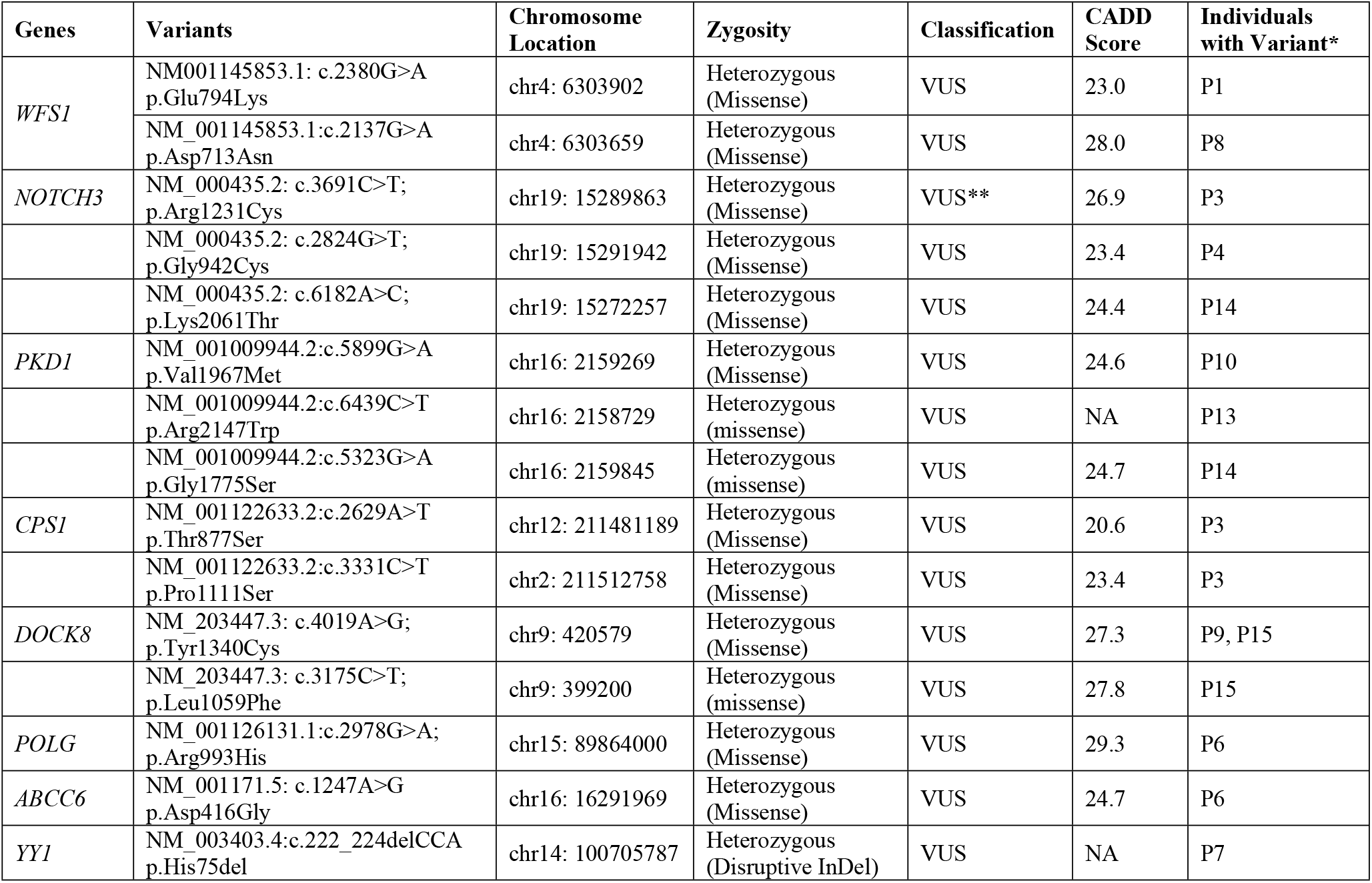
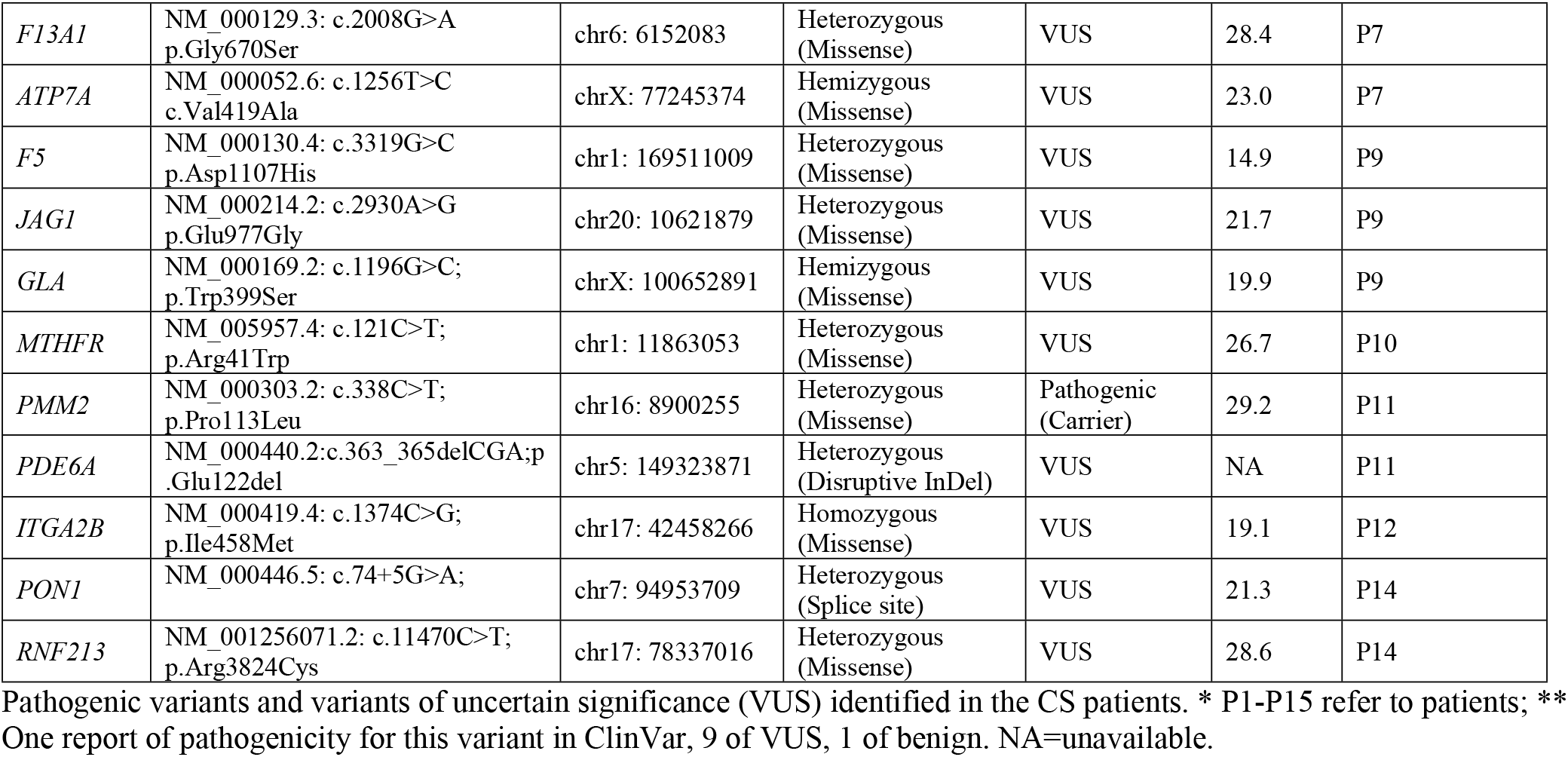
Variants identified in the CS patients. Pathogenic variants and variants of uncertain significance (VUS) identified in the CS patients.

| Genes | Variants | Chromosome Location | Zygosity | Classification | CADD Score | Individuals with Variant* |
| --- | --- | --- | --- | --- | --- | --- |
| *WFS1* | NM001145853.1: c.2380G>A<br>p.Glu794Lys | chr4: 6303902 | Heterozygous (Missense) | VUS | 23.0 | P1 |
|  | NM_001145853.1: c.2137G>A<br>p.Asp713Asn | chr4: 6303659 | Heterozygous (Missense) | VUS | 28.0 | P8 |
| *NOTCH3* | NM_000435.2: c.3691C>T;<br>p.Arg1231Cys | chr19: 15289863 | Heterozygous (Missense) | VUS** | 26.9 | P3 |
|  | NM_000435.2: c.2824G>T;<br>p.Gly942Cys | chr19: 15291942 | Heterozygous (Missense) | VUS | 23.4 | P4 |
|  | NM_000435.2: c.6182A>C;<br>p.Lys2061Thr | chr19: 15272257 | Heterozygous (Missense) | VUS | 24.4 | P14 |
| *PKD1* | NM_001009944.2: c.5899G>A<br>p.Val1967Met | chr16: 2159269 | Heterozygous (Missense) | VUS | 24.6 | P10 |
|  | NM_001009944.2: c.6439C>T<br>p.Arg2147Trp | chr16: 2158729 | Heterozygous (missense) | VUS | NA | P13 |
|  | NM_001009944.2: c.5323G>A<br>p.Gly1775Ser | chr16: 2159845 | Heterozygous (missense) | VUS | 24.7 | P14 |
| *CPS1* | NM_001122633.2: c.2629A>T<br>p.Thr877Ser | chr12: 211481189 | Heterozygous (Missense) | VUS | 20.6 | P3 |
|  | NM_001122633.2: c.3331C>T<br>p.Pro1111Ser | chr2: 211512758 | Heterozygous (Missense) | VUS | 23.4 | P3 |
| *DOCK8* | NM_203447.3: c.4019A>G;<br>p.Tyr1340Cys | chr9: 420579 | Heterozygous (Missense) | VUS | 27.3 | P9, P15 |
|  | NM_203447.3: c.3175C>T;<br>p.Leu1059Phe | chr9: 399200 | Heterozygous (missense) | VUS | 27.8 | P15 |
| *POLG* | NM_001126131.1: c.2978G>A;<br>p.Arg993His | chr15: 89864000 | Heterozygous (Missense) | VUS | 29.3 | P6 |
| *ABCC6* | NM_001171.5: c.1247A>G<br>p.Asp416Gly | chr16: 16291969 | Heterozygous (Missense) | VUS | 24.7 | P6 |
| *YY1* | NM_003403.4: c.222_224delCCA<br>p.His75del | chr14: 100705787 | Heterozygous (Disruptive InDel) | VUS | NA | P7 |
| *F13A1* | NM_000129.3: c.2008G>A<br>p.Gly670Ser | chr6: 6152083 | Heterozygous<br>(Missense) | VUS | 28.4 | P7 |
| *ATP7A* | NM_000052.6: c.1256T>C<br>c.Val419Ala | chrX: 77245374 | Hemizygous<br>(Missense) | VUS | 23.0 | P7 |
| *F5* | NM_000130.4: c.3319G>C<br>p.Asp1107His | chr1: 169511009 | Heterozygous<br>(Missense) | VUS | 14.9 | P9 |
| *JAG1* | NM_000214.2: c.2930A>G<br>p.Glu977Gly | chr20: 10621879 | Heterozygous<br>(Missense) | VUS | 21.7 | P9 |
| *GLA* | NM_000169.2: c.1196G>C;<br>p.Trp399Ser | chrX: 100652891 | Hemizygous<br>(Missense) | VUS | 19.9 | P9 |
| *MTHFR* | NM_005957.4: c.121C>T;<br>p.Arg41Trp | chr1: 11863053 | Heterozygous<br>(Missense) | VUS | 26.7 | P10 |
| *PMM2* | NM_000303.2: c.338C>T;<br>p.Pro113Leu | chr16: 8900255 | Heterozygous<br>(Missense) | Pathogenic<br>(Carrier) | 29.2 | P11 |
| *PDE6A* | NM_000440.2:c.363_365delCGA;p<br>.Glu122del | chr5: 149323871 | Heterozygous<br>(Disruptive InDel) | VUS | NA | P11 |
| *ITGA2B* | NM_000419.4: c.1374C>G;<br>p.Ile458Met | chr17: 42458266 | Homozygous<br>(Missense) | VUS | 19.1 | P12 |
| *PON1* | NM_000446.5: c.74+5G>A; | chr7: 94953709 | Heterozygous<br>(Splice site) | VUS | 21.3 | P14 |
| *RNF213* | NM_001256071.2: c.11470C>T;<br>p.Arg3824Cys | chr17: 78337016 | Heterozygous<br>(Missense) | VUS | 28.6 | P14 |

Pathogenic variants and variants of uncertain significance (VUS) identified in the CS patients. \* P1-P15 refer to patients; \*\* One report of pathogenicity for this variant in ClinVar, 9 of VUS, 1 of benign. NA=unavailable.

### Gene set enrichment and STRING analyses

Putative deleterious variants could contribute to genetic risk either as complex heterozygotes in a single gene (e.g. Table 1 P3 at *CPS1*; P15 at *DOCK8*) or at interacting proteins in common pathways. To determine how proteins encoded by genes carrying putative deleterious variants might interact we carried out gene-set enrichment analysis in EnrichR using the 19 genes carrying putative deleterious variants (Table 2). Enrichr showed significant enrichment for gene-sets in phenotypes (e.g. hemorrhage; abnormal blood coagulation), pathways (e.g. common pathway of fibrin clot formation; platelet degranulation; response to elevated platelet cytosolic Ca^2+^), and cellular components (e.g. platelet alpha granule) that could contribute to CS. STRING analysis demonstrated that 6 of the genes common across enriched gene-sets, *IGA2B, F13A1, F5, ATP7A, GLA* and *ABCC6* interact at the protein level (S1 Fig). This provides provisional functional support that patients who carry putative deleterious variants at more than one of these genes, e.g. P7 with variants at *F13A1* and *ATP7A*, could be investigated for polygenic inheritance involving these genes. Indeed, multiple patients carried putative deleterious variants across more than one gene in these enriched gene-sets (Table 1).

**Table 2.** Enrichr analysis. Results of Enrichr analysis (P<0.001; adjusted P-values <0.01) for gene sets identified from 19 genes carrying 26 putative deleterious VUS (missense CADD score ≥15 or in-frame deletions) in patients with cryptogenic stroke.

| Table and Term | Overlap | P-value | Odds Ratio | Combined Score | Genes |
| --- | --- | --- | --- | --- | --- |
| **MGI Mammalian Phenotype Level 4** |  |  |  |  |  |
| MP:0001914 Hemorrhage | 7 of 325 | 1.20E-08 | 36.1 | 658 | *JAG1;PMM2;ITGA2B;F13A1;ATP7A;PKD1;F5* |
| MP:0002551 Abnormal Blood Coagulation | 3 of 51 | 1.47E-05 | 77.8 | 866 | *ITGA2B;F13A1;F5* |
| MP:0006278 Aortic Aneurysm | 2 of 8 | 2.39E-05 | 391.7 | 4168 | *ATP7A;PKD1* |
| MP:0031152 Subcutaneous Hemorrhage | 2 of 9 | 3.07E-05 | 335.7 | 3489 | *ITGA2B;F13A1* |
| MP:0005435/5243 Hemoperitoneum/Hemothorax | 2 of 10 | 3.83E-05 | 293.7 | 2987 | *F13A1;ATP7A* |
| MP:0004363 Stria Vascularis Degeneration | 2 of 13 | 6.63E-05 | 213.6 | 2055 | *WFS1;POLG* |
| MP:0010574 Dilated Aorta | 2 of 15 | 8.91E-05 | 180.7 | 1685 | *ATP7A;GLA* |
| MP:0003674 Oxidative Stress | 3 of 127 | 2.25E-04 | 30.0 | 252 | *PON1;ATP7A;POLG* |
| MP:0002833 Increased Heart Weight | 4 of 360 | 3.23E-04 | 14.7 | 118 | *NOTCH3;PKD1;GLA;POLG* |
| **Go Cellular component 2023** |  |  |  |  |  |
| Platelet Alpha Granule (GO:0031091) | 3 of 89 | 7.84E-05 | 43.4 | 410 | *ITGA2B;F13A1;F5* |
| **Jensen Compartments** |  |  |  |  |  |
| Platelet alpha granule | 3 of 72 | 4.16E-05 | 54.1 | 546 | *ITGA2B;F13A1;F5* |
| **Jensen Diseases** |  |  |  |  |  |
| Cerebrovascular disease | 6 of 225 | 4.55E-08 | 41.6 | 704 | *NOTCH3;ITGA2B;PON1;MTHFR;GLA;F5* |
| Atherosclerosis | 3 of 52 | 1.56E-05 | 76.3 | 884 | *RNF213;CPS1;MTHFR* |
| Cadasil | 2 of 9 | 3.07E-05 | 335.7 | 3489 | *NOTCH3;JAG1* |
| **Reactome 2022** |  |  |  |  |  |
| R-HSA-114608 Platelet Degranulation | 3 of 128 | 2.30E-04 | 29.7 | 249 | *ITGA2B;F13A1;F5* |
| R-HSA-140875 Common Pathway of Fibrin Clot Formation | 2 of 22 | 1.95E-04 | 117.46 | 1003 | *F13A1;F5* |
| R-HSA-76005 Response to Elevated Platelet Cytosolic Ca<sup>2+</sup> | 3 of 133 | 2.58E-04 | 28.6 | 344237 | *ITGA2B;F13A1;F5* |
| R-HSA-9013507 NOTCH3 Activation | 2 of 25 | 2.53E-04 | 102.1 | 845 | *NOTCH3;JAG1* |
| R-HSA-140877 Formation of Fibrin Clot (Clotting Cascade) | 2 of 39 | 6.20E-04 | 63.4 | 468 | *F13A1;F5* |

| **Human Phenotype Ontology** |  |  |  |  |  |
| --- | --- | --- | --- | --- | --- |
| Stroke (HP:0001297) | 8 of 33 | 1.63E-18 | 580.5 | 23776 | *NOTCH3;JAG1;CPS1;WFS1;ABCC6;PMM2;MTHFR;GLA* |
| Autosomal dominant inheritance (HP:0000006) | 10 of 1134 | 1.91E-08 | 18.6 | 331 | *NOTCH3;JAG1;WFS1;DOCK8;ABCC6;ITGA2B;PON1;PKD1;F5;POLG* |
| Autosomal recessive inheritance (HP:0000007) | 11 of 1722 | 7.37E-08 | 14.7 | 241 | *CPS1;WFS1;DOCK8;ABCC6;PMM2;ITGA2B;F13A1;MTHFR;PDE6A;F5;POLG* |
| Aneurysm (HP:0002617) | 3 of 39 | 6.50E-06 | 103.8 | 1241 | *ABCC6;ATP7A;PKD1* |
| Myocardial infarction (HP:0001658) | 2 of 11 | 4.68E-05 | 261.1 | 2603 | *ABCC6;GLA* |
| Abnormality of the common coagulation pathway (HP:0010990) | 2 of 17 | 1.15E-04 | 156.6 | 1420 | *F13A1; F5* |
| Congestive heart failure (HP:0001635) | 3 of 128 | 2.30E-04 | 29.8 | 250 | *WFS1;ABCC6;GLA* |

## Discussion

We identified 26 putative deleterious VUS in Indian CS patients in genes previously associated with stroke or stroke-related clinical phenotypes. While none were known pathogenic variants in ClinVar, all have the potential to contribute to clinical CS if confirmed as functionally deleterious. Firstly, three patients were either homozygous (P12 at *ITGA2B*) or hemizygous (P7 at *ATP7A*; P9 at *GLA*) for VUS which could be directly disease causing. *ITGA2B* is associated with ischemic stroke with its expression in COVID-19-related stroke related to elevated platelet cytosolic Ca^2+^ [7]. *ATP7A* encodes a copper-transporting ATPase that predicts cuproptosis, a novel form of programmed cell death in ischemic stroke [8]. P7 also carried a VUS at *F13A1* encoding coagulation factor XIII, a key differentially expressed gene associated with ischemic stroke [9], and a disruptive InDel in *YY1* involved in stroke induction [10]. The combination of VUS at *ATP7A* and *F13A1* is interesting given STRING evidence for protein interaction. Exome-based analysis identified two pathogenic variants in *GLA* in 172 ischemic stroke patients [11]. P9 also carried VUS at *F5, DOCK8* and *JAG1*. Central nervous system vasculitis and stroke are complications of *DOCK8* deficiency [12], *F5* encoding coagulation factor V is a novel biomarker in ischemic stroke [13], while *JAG1* encodes Notch receptor ligand jagged-1 (cf. below).

Compound heterozygosity at a single gene could also increase disease risk, e.g. P15 compound heterozygote at *DOCK8*, a gene associated with stroke [12]. P3 was compound heterozygote at *CPS1*, a gene regulating heritable circulating homocysteine levels associated with stroke [14]. P3 also carried a *NOTCH3* VUS, pathogenic variants at which were also identified exome-based analysis of juvenile ischemic stroke [11]. P14 carried variants at *NOTCH3, PKD1, PON1* and *RNF213*. A blended phenotype of variants at *NOTCH3* and *RNF213* is associated with temporal pole infarcts in stroke episodes [15]. Inhibition of the BCL6/miR-31/PKD1 axis attenuates oxidative stress-induced neuronal damage and reduces cerebral infarction [16]. P10 and P13 carried different VUS at *PKD1*, that in P10 in combination with a VUS at *MTHFR*. A functional variant at *MTHFR* downregulates methylenetetrahydrofolate reductase activity and is associated with increased risk of small vessel stroke [17]. P14 carried a third *PKD1* variant in combination with a heterozygous splice site variant at *PON1* that encodes paraoxonase 1 which is protective in ischemic stroke [18].

Additional CS patients were heterozygous for putative deleterious variants that could be dominant or contribute to polygenic inheritance. P1 and P8 each carried a single VUS at *WFS1*. Deleterious variants at *WFS1* cause Wolfram syndrome 1 (OMIM **#**222300), which can be associated with stroke-like clinical signs. P11 was heterozygous for a disruptive InDel in the phosphodiesterase-6A gene *PDE6A*, causative for retinitis pigmentosa 43 (OMIM *613810). P11 was also a carrier of a known pathogenic variant at *PMM2*. Pathogenic variants in *PMM2* cause congenital disorder of glycosylation, type Ia (OMIM **#**212065) with stroke-like episodes. Although normally observed on the background of severe developmental delay, ataxia and dysmorphism, the disorder has been identified in a developmentally normal young individual presenting with isolated stroke-like episodes [19]. P6 carried VUS at both *POLG* and *ABCC6*. DNA polymerase gamma encoded by *POLG* plays a role in the replication of mitochondrial DNA with pathogenic variants associated with mitochondrial disorders (OMIM **\***174763) that can include stroke-like episodes. *ABCC6* belongs to a family of ATP-binding cassette transmembrane transporters, with pathogenic variants also associated with disorders (OMIM **\***603234) that include arterial calcification and myocardial infarction. In a study of mutations in Mendelian stroke genes in 1,033 early onset stroke patients, clinically relevant VUS were identified at *ABCC6* (n=53), *RNF213* (n=59) and *NOTCH3* (n=15) [20], indicating that variants at all of these genes are worthy of further investigation in our CS patient group.

Here we identified 26 potentially clinically relevant VUS in 19 genes in 16 CS patients for whom follow-up extended family studies and/or functional gene-editing laboratory investigations are warranted. These genes could become important targets for development of biomarkers of CS and therapeutic interventions.

## Acknowledgements

We would like to thank Dr. V N Mishra, Dr. Deepika Joshi, Dr. R N Chaurasia, Dr. Varun Kumar Singh, and Dr. Anand Kumar for their support in the study, and BHU for their infrastructural support.

## Authors’ contributions

Study design, funding and supervision: AP. Data collection and analysis: PD, VNM, RK, and JMB. Manuscript writing and editing: PD, RK and JMB. All authors have read and approved the manuscript.

## Data Availability Statement

All data relevant to results of this study, including generated and analysed data, are contained within this article and the supporting files.

## Supporting information

**S1 Fig**. Results of STRING analysis for 19 genes with VUS in CS patients. Nodes (= query proteins) are represented by coloured circles; filled nodes indicate that the protein structure is known or predicted. Protein-protein interactions are represented by coloured lines as indicated in the key. These interactions indicated that proteins contribute to a shared function but does not necessarily mean they are physically binding each other (https://string-db.org/).

**S1 Table** Panel of 220 stroke-related genes included during targeted analysis of exome sequencing data.

**S2 Table** Characteristics, clinical data and family history for stroke and stroke-related risk factors for CS patients.

## References

1. Salomi BSB, Solomon R, Turaka VP, Aaron S, Christudass CS. Cryptogenic Stroke in the Young: Role of Candidate Gene Polymorphisms in Indian Patients with Ischemic Etiology. Neurol India. 2021;69(6):1655–62. Epub 2022/01/05. doi: 10.4103/0028-3886.333441. PubMed PMID: 34979665.

2. Bevan S, Traylor M, Adib-Samii P, Malik R, Paul NL, Jackson C, et al. Genetic heritability of ischemic stroke and the contribution of previously reported candidate gene and genomewide associations. Stroke. 2012;43(12):3161–7. Epub 2012/10/09. doi: 10.1161/STROKEAHA.112.665760. PubMed PMID: 23042660.

3. Kumar A, Chauhan G, Sharma S, Dabla S, Sylaja PN, Chaudhary N, et al. Association of SUMOylation Pathway Genes With Stroke in a Genome-Wide Association Study in India. Neurology. 2021;97(4):e345–e56. Epub 2021/05/26. doi: 10.1212/WNL.0000000000012258. PubMed PMID: 34031191; PubMed Central PMCID: PMCPMC8362360.

4. Bomba L, Walter K, Soranzo N. The impact of rare and low-frequency genetic variants in common disease. Genome Biol. 2017;18(1):77. Epub 2017/04/30. doi: 10.1186/s13059-017-1212-4. PubMed PMID: 28449691; PubMed Central PMCID: PMCPMC5408830.

5. Adams HP, Jr., Bendixen BH, Kappelle LJ, Biller J, Love BB, Gordon DL, et al. Classification of subtype of acute ischemic stroke. Definitions for use in a multicenter clinical trial. TOAST. Trial of Org 10172 in Acute Stroke Treatment. Stroke. 1993;24(1):35–41. Epub 1993/01/01. doi: 10.1161/01.str.24.1.35. PubMed PMID: 7678184.

6. Kircher M, Witten DM, Jain P, O’Roak BJ, Cooper GM, Shendure J. A general framework for estimating the relative pathogenicity of human genetic variants. Nat Genet. 2014;46(3):310–5. doi: 10.1038/ng.2892. PubMed PMID: 24487276; PubMed Central PMCID: PMCPMC3992975.

7. Cen G, Liu L, Wang J, Wang X, Chen S, Song Y, et al. Weighted Gene Co-Expression Network Analysis to Identify Potential Biological Processes and Key Genes in COVID-19-Related Stroke. Oxid Med Cell Longev. 2022;2022:4526022. Epub 2022/05/14. doi: 10.1155/2022/4526022. PubMed PMID: 35557984; PubMed Central PMCID: PMCPMC9088964 publication of this article.

8. Fan X, Chen H, Jiang F, Xu C, Wang Y, Wang H, et al. Comprehensive analysis of cuproptosis-related genes in immune infiltration in ischemic stroke. Front Neurol. 2022;13:1077178. Epub 2023/02/24. doi: 10.3389/fneur.2022.1077178. PubMed PMID: 36818726; PubMed Central PMCID: PMCPMC9933552.

9. Lin L, Guo C, Jin H, Huang H, Luo F, Wang Y, et al. Integrative multi-omics approach using random forest and artificial neural network models for early diagnosis and immune infiltration characterization in ischemic stroke. Front Neurol. 2024;15:1475582. Epub 2024/12/19. doi: 10.3389/fneur.2024.1475582. PubMed PMID: 39697434; PubMed Central PMCID: PMCPMC11652371.

10. Pabian-Jewula S, Bragiel-Pieczonka A, Rylski M. Ying Yang 1 engagement in brain pathology. J Neurochem. 2022;161(3):236–53. Epub 2022/02/25. doi: 10.1111/jnc.15594. PubMed PMID: 35199341.

11. Hartl J, Hartberger J, Wunderlich S, Cordts I, Bafligil C, Sturm M, et al. Exome-based gene panel analysis in a cohort of acute juvenile ischemic stroke patients:relevance of NOTCH3 and GLA variants. J Neurol. 2023;270(3):1501–11. Epub 2022/11/22. doi: 10.1007/s00415-022-11401-7. PubMed PMID: 36411388; PubMed Central PMCID: PMCPMC9971083.

12. AlKhater SA. CNS vasculitis and stroke as a complication of DOCK8 deficiency: a case report. BMC Neurol. 2016;16:54. Epub 2016/04/27. doi: 10.1186/s12883-016-0578-3. PubMed PMID: 27113444; PubMed Central PMCID: PMCPMC4845487.

13. Liu J, Si Z, Liu J, Zhang X, Xie C, Zhao W, et al. Machine learning identifies novel coagulation genes as diagnostic and immunological biomarkers in ischemic stroke. Aging (Albany NY). 2024;16(7):6314–33. Epub 2024/04/05. doi: 10.18632/aging.205706. PubMed PMID: 38575196; PubMed Central PMCID: PMCPMC11042924.

14. Williams SR, Yang Q, Chen F, Liu X, Keene KL, Jacques P, et al. Genome-wide meta-analysis of homocysteine and methionine metabolism identifies five one carbon metabolism loci and a novel association of ALDH1L1 with ischemic stroke. PLoS Genet. 2014;10(3):e1004214. Epub 2014/03/22. doi: 10.1371/journal.pgen.1004214. PubMed PMID: 24651765; PubMed Central PMCID: PMCPMC3961178.

15. Saito S, Hosoki S, Yamaguchi E, Ishiyama H, Abe S, Yoshimoto T, et al. Blended Phenotype of NOTCH3 and RNF213 Variants With Accelerated Large and Small Artery Crosstalk: A Case Report and Literature Review. Neurol Genet. 2024;10(5):e200176. Epub 2024/09/11. doi: 10.1212/NXG.0000000000200176. PubMed PMID: 39257469; PubMed Central PMCID: PMCPMC11384340 disclosures.

16. Wei P, Chen H, Lin B, Du T, Liu G, He J, et al. Inhibition of the BCL6/miR-31/PKD1 axis attenuates oxidative stress-induced neuronal damage. Exp Neurol. 2021;335:113528. Epub 2020/11/16. doi: 10.1016/j.expneurol.2020.113528. PubMed PMID: 33189730.

17. Wang M, Daghlas I, Zhang Z, Gill D, Liu D. MTHFR Polymorphisms, Homocysteine Elevation, and Ischemic Stroke Susceptibility in East Asian and European Populations. Neurology. 2025;104(3):e210245. Epub 2025/01/09. doi: 10.1212/WNL.0000000000210245. PubMed PMID: 39787475.

18. Asahara N, Ebisu H, Yuki S, Fujita R, Kojima S. Paraoxonase 1 ameliorates neurological symptoms and motor coordination impairment caused by cerebral ischemia-reperfusion injury. Biomed Pharmacother. 2024;182:117792. Epub 2024/12/30. doi: 10.1016/j.biopha.2024.117792. PubMed PMID: 39733589.

19. Farmania R, Jain P, Sharma S, Aneja S. Unusual Presentation of PMM2-Congenital Disorder of Glycosylation With Isolated Strokelike Episodes in a Young Girl. Journal of child neurology. 2019;34(7):410–4. Epub 2019/03/13. doi: 10.1177/0883073819833543. PubMed PMID: 30857461.

20. Park HK, Lee KJ, Park JM, Kang K, Lee SJ, Kim JG, et al. Prevalence of Mutations in Mendelian Stroke Genes in Early Onset Stroke Patients. Ann Neurol. 2023;93(4):768–82. Epub 2022/12/22. doi: 10.1002/ana.26575. PubMed PMID: 36541592.

